# Strengthening a collaborative approach to implementing surveillance systems: Lessons from the Enhanced Gonococcal Antimicrobial Surveillance Programme (EGASP) in Malawi and Zimbabwe

**DOI:** 10.1101/2024.03.26.24304934

**Authors:** Phiona Vumbugwa, Ismael Maatouk, Anna Machiha, Mitch Matoga, Collins Mitambo, Rose Nyirenda, Ishmael Nyasulu, Muchaneta Mugabe, Mkhokheli Ngwenya, Yamuna Mundade, Teodora Wi, Magnus Unemo, Olusegun O. Soge

## Abstract

**Background:** With the number of antimicrobials available to effectively treat gonorrhoea rapidly diminishing, surveillance of antimicrobial–resistant *Neisseria gonorrhoeae* (NG) is critical for global public health security activity. Many low-and-middle-income countries (LMICs) have gaps in their existing sexually transmitted infections (STIs) surveillance systems that negatively impact global efforts geared towards achieving the United Nations (UN) Sustainable Development Goals (SDGs). This paper explains the contribution of collaborative surveillance systems to health systems strengthening (HSS) learning from integrating NG surveillance into existing Ministries of Health’s (MoH) antimicrobial resistance (AMR) surveillance services in Malawi and Zimbabwe.

**Methods:** We used the WHO Enhanced Gonococcal Antimicrobial Surveillance Programme (EGASP) implementation experiences in Malawi and Zimbabwe to demonstrate the collaboration in AMR and STI surveillance. We conducted qualitative interviews with purposively selected health managers directly participating in the AMR and STI programs using a standardized key informant guide to describe how to plan for a collaborative surveillance system. Qualitative thematic analysis was conducted to delineate stakeholders’ recommendations using the health systems’ building blocks.

**Results:** Stakeholder engagement, prioritization of needs, and power to negotiate were key drivers to a successful collaborative surveillance system. Weak governance, policies, lack of accountability, and different priorities, coupled with weak collaborations, workforce, and health information systems, were challenges faced in having effective collaborative surveillance systems. Data availability, use, and negotiation power were key drivers for the prioritization of collaborative surveillance. Including collaborative surveillance in primary health services and increasing government budget allocation for surveillance were recommended.

**Conclusions:** Strengthening collaborative surveillance systems in LMICs using a people-centered approach increases transparency and accountability and empowers national institutions, communities, and stakeholders to engage in sustainable activities that potentially strengthen health systems. EGASP implementations in Zimbabwe and Malawi serve as models for other countries planning to implement or improve collaborative surveillance systems in their context.

## Introduction

In 2020, the World Health Organization (WHO) estimated 82.4 million new global infections with Neisseria gonorrhoeae (NG) in adults aged 15 to 49 years, with the highest prevalence in low-and-middle-income countries (LMICs) (1–3). Globally, many countries have suboptimal sexually transmitted infections (STIs) surveillance systems and laboratory diagnostics because of limited epidemiologic and laboratory capacity, lack of political will, and funding (4,5). Lack of antimicrobial resistance (AMR) surveillance further threatens the progress of achieving the United Nations’ (UN) Sustainable Development Goals (SDGs), which aim to ensure access to safe and affordable medicines and vaccines for all (6–8). Health systems are weakened by a high disease burden, threatening the countries’ economies as productive populations spend more time in hospitals (9). Most Sub-Saharan African (SSA) countries rely on syndromic management of STIs due to limited access to diagnostic assays (10–12). There is a growing need for countries to implement collaborative STI surveillance to provide effective treatment.

There is limited data in countries to support the review of STI treatment guidelines. Few countries conduct regular treatment performance assessments, which require laboratory diagnostics (13). Countries’ surveillance systems implementation is guided by National Action Plans for Health Security (NAPHS). Progress is limited by technical, operational, financial, and political challenges (14). While funding mechanisms like The Pandemic Fund and the Global Fund COVID-19 Response Mechanism provided new opportunities for countries to prioritize and improve epidemic preparedness, effectively leveraging funding can be challenging (4,15). Focusing on collaborative surveillance would inform changes needed to respond to and manage AMR and strengthen health systems.

Understanding the process of planning collaborative surveillance potentiates health leaders to make appropriate decisions for better population health outcomes. The WHO Enhanced Gonococcal Antimicrobial Surveillance Programme (EGASP) is a special project under the Global Antimicrobial Resistance and Use Surveillance System (GLASS) to strengthen sentinel surveillance for gonococcal AMR (16). EGASP is a collaborative effort between the WHO, the United States Centers for Disease Control and Prevention (CDC), and WHO collaborating centers on STIs. The EGASP enhances the laboratory capacity in early detection and reporting of NG strains with elevated minimum inhibitory concentrations (MICs) to the recommended treatment for gonorrhoea and emerging resistance to new antibiotics for gonorrhoea at the national level (17,18). Malawi and Zimbabwe are amongst the 13 initial EGASP countries and implemented EGASP in 2023 through their national AMR surveillance programs. The global AMR for NG information is reported and shared through GLASS IT WHONET, a Windows-based digital health information system (DHIS) database that captures, analyzes, and manages microbiology laboratory data (19). Collaborative surveillance is a multifaceted concept crucial in enhancing security, safety, and efficiency across all the pillars of health systems strengthening (HSS).

Literature has revealed that several organizations have integrated various programs and projects. There is little documented evidence on collaborative surveillance, and many organizations struggle to understand the best way possible (20). Understanding the considerations for implementing collaborative surveillance supports the implementation of the NAPHS strategic toolkit, which shows promises to strengthen health systems. Learning from the EGASP implementation in Malawi and Zimbabwe, the present study details key considerations when implementing collaborative surveillance. Using the six health systems building blocks, we describe challenges to prioritizing collaborative surveillance systems and recommendations when collaborating surveillance systems to contribute to health system strengthening.

## Methodology

### Design

The study follows the qualitative case study design where key informant interviews (KIIs) were conducted using a structured interview guide. Drawing experiences from the EGASP in Malawi and Zimbabwe settings, we demonstrate how collaborative surveillance was planned, the challenges faced, and recommendations for future countries planning to implement collaborative surveillance in routine settings. The study was conducted from April 1^st^ to November 29^th^ 2023.

### Setting /context

Malawi and Zimbabwe are landlocked countries proximal to each other located in the Southern region of Africa. The two countries were selected for EGASP implementation based on their burden of STIs and prior participation in the WHO GASP in 2017. Both countries have functioning microbiology reference laboratories that can perform culture, antimicrobial susceptibility testing, and quality assurance measures. The availability of a clear leadership structure that supports, manages, and guides the implementation of the STI and AMR supported the establishment of sentinel sites required to implement the EGASP in both countries. The EGASP program is run at high-volume STI sites where clinicians systematically collect swabs from men attending the clinic with a suspected urogenital gonorrhoea episode (such as the presence of urethral discharge). The urethral specimens are sent to a reference lab for NG culture and antimicrobial susceptibility testing (AST). An implementing partner, the University of North Carolina (UNC) Project Malawi Research Institute in Malawi and the government-owned National Microbiology Reference Laboratory (NMRL) in Zimbabwe support the EGASP NG lab activities.

### Participants selection, recruitment, and eligibility

Eight health managers directly participating in the AMR, STIs, and EGASP surveillance systems, working in the Ministry of Health (MoH) and its partners at the national level, were purposely selected. All participants were leaders in a clinical or laboratory decision-making role with experience in leading, coordinating, implementing, or providing technical support to the STIs and AMR programs. Only participants involved in the EGASP with a leadership role who consented to be interviewed were included.

### Health systems building blocks framework

Using the World Health Organization’s (WHO) six health systems building blocks framework, participants gave their perspectives on challenges and key considerations when prioritizing an expanded scope of collaborative routine surveillance. The framework is used in public health to guide strengthening health systems and improve healthcare delivery, describing the essential components that, when properly developed and integrated, can enhance healthcare systems’ overall resilience and effectiveness (13). Table 1 below shows the six building blocks and their focus on collaborative surveillance.

**Table 1:** Health systems strengthening framework in the context of WHO EGASP.

| Health systems building block | Focus | Areas to strengthen |
| --- | --- | --- |
| **Leadership and Governance** | Overall coordination and management of EGASP | Governance structures, regulations, and policies that guide surveillance system functioning |
| **Information Systems** | Collecting, managing, and utilizing data to inform decision-making and monitor the health status of populations. | Timely and quality data collection, analysis, use, and dissemination. |
| **Health Financing** | Adequate and sustainable financing of EGASP | The mobilization, allocation, and utilization of financial resources for the surveillance system |
| **Health Workforce** | Skilled and motivated workforce to implement EGASP | Training, recruitment, retention, and deployment of healthcare professionals within the surveillance system |
| **Service delivery** | Availability and accessibility of essential healthcare services | Improving the quality of healthcare services makes them more responsive to community needs (NG treatment). |
| **Medical & laboratory supplies** | Access to essential medical products and technologies | Procurement, supply chain management, and quality assurance of these products. |

**Table 2:** Participants profile.

| Organization | National Level | Above national level | Study Identification |
| --- | --- | --- | --- |
| Ministry of Health | 5 | 0 | MoH |
| Funding Agent | 0 | 1 | FA |
| Implementing Partners | 1 | 1 | IP |
| Total | 6 | 2 |  |

### Data collection

Participants were identified during the EGASP laboratory and sentinel site assessment visits in Malawi and Zimbabwe, i.e., while providing technical support to country leadership. Based on the participants’ availability, interviews were scheduled for 30 minutes, conducted, and recorded via ZOOM. Participants were emailed the key informant guide three to seven days before the interviews. Interviews were conducted by the lead author in English. We reviewed grey literature from reports compiled from EGASP country visits, countries’ sentinel site assessment reports, discussions from implementation meetings, and documents shared by participants on NG surveillance to augment interview data.

### Data analysis

The lead author manually transcribed and analyzed all recordings using Dedoose version 9.0. The transcription process involved reading the text multiple times, thus enhancing the author’s familiarity with the transcripts. The analysis was done per each health system building block. Researchers went on to code every line of the transcripts to identify emergent themes. This was followed by a grouping of codes with similar ideas. To better understand key informant’s accounts, we used the notes and documents referenced by participants collected during the interviews and country visits. A descriptive analysis detailed the challenges, facilitators, and recommendations in prioritizing collaborative surveillance using the health systems’ building blocks.

### Positionality statement

The professional knowledge in healthcare and practice-based experience from the EGASP that the lead analyst (PV) participated in helped to conceptualize the research. Participants were engaged in a sensitive and open manner. The research was conducted ethically, respecting inclusive and equitable workspace for all gender identities.

### Ethical considerations

The University of Washington (UW) internal review board (IRB) approved the study as non-human subject under the study number UW IRB STUDY00018156. The research paper was reviewed by WHO publication clearance committee. All participants gave informed verbal consent to participate and record at the start of the interviews. Verbal consent was documented on the a table in the interview script. No personally identifiable information (PII) was collected. The data was stored in a secure SharePoint folder on a computer with a protected password. This data is available upon request.

## Results

### Participants profile

Of the six participants from the national level, three were from each country. Five participants identified themselves as males, two as females, and one preferred not to disclose their gender identity. All participants supported the EGASP implementation in Malawi and Zimbabwe.

### Key considerations when collaborating on a surveillance system

Participants highlighted using existing surveillance priorities, strategies, resources, and data as considerations countries use to plan an effective collaborative surveillance system. When planning for the EGASP implementation, health leaders from the national level indicated that they used lessons learned from Integrated Disease Surveillance Response (IDSR). However, this is activated in emergencies. Key lessons from the EGASP collaborative surveillance included using existing structures, people, processes, resources, coordinated plans, data, programs, and involvement in STI and NG surveillance implementation.

A national leader from Malawi indicated the need to know the landscape, programs available, stakeholders involved, and the flexibility of the programs for easy and efficient collaboration. In support, participants stated that for EGASP in Malawi, the leadership team used the existing collaborations to support the uptake and implementation of surveillance. Previously, the UNC Project Malawi research institute, having a private laboratory fully equipped to run a national surveillance system, worked with the MoH in Malawi in conducting NG resistance surveillance for gentamicin, the first-line drug for syndromic treatment of urethral discharge in the country. In collaboration with the laboratory, sentinel sites run within the MoH structures with dedicated health workers responsible for data and sample collection. The sentinel site has a small laboratory for specimen handling, packaging, and shipment to the reference laboratory. During working hours, a dedicated courier transports specimens on a motorcycle from the clinic site to the reference laboratory. The UNC Project Malawi Research Institute also strengthened MoH’s national and regional laboratory reference centers, conducting the AMR surveillance, including quality sample collection, handling, and processing, which resulted in the two national laboratories becoming ISO 15189 certified. However, the country’s public health system faces human resources shortages in clinical and laboratory systems. UNC Project Malawi, therefore, was selected to start the implementation of EGASP in Malawi while building the MoH capacity for EGASP.

A participant from Zimbabwe emphasized that planning collaborative surveillance without engaging relevant stakeholders would stall uptake and impede implementation. Another participant cited that using existing structures and systems, low-hanging fruits, evidence, and experience from other public health programs was essential in sustainable collaborative surveillance. The participants stated that they used the expertise from implementing WHO GASP in 2018 to plan the WHO EGASP 2023 implementation. Three sentinel sites were selected, with the National Microbiology Reference Laboratory (NMRL) conducting the EGASP laboratory activities, including NG culture and AST. Although the sentinel sites chosen were located far from the NMRL, the district laboratories close to the facilities had personnel knowledgeable about preserving the specimens before transportation.

All participants agreed that having a solid coordination system and a clearly defined structure simplifies stakeholder communication and reporting while implementing the EGASP surveillance system. One participant cited their country having rekindled collaboration with the neighboring country, South Africa, in coordinating support with procurement and supply for critical reagents and laboratory supplies that are not readily available in the country. Implementation was through the MoH structures for both countries, with technical support from the WHO and WHO collaborating centers in the USA and Sweden.

### Governance and Leadership

Respondents cited policies required to prioritize collaborative surveillance, including the National Health Strategic Plan, Joint Midterm Reviews, the Diagnostic Network Plan for public health surveillance, laboratory preparedness, and response, as one of the ten global strategic focus areas. Other frameworks to utilize were One Health Framework for Africa and the Biosafety and Biosecurity Initiative launched in 2019, as one participant stated:

“*It is easier for a leader to institute workforce laws backed by policies and laws*.” *MoH*

Several participants agreed that policies empower leaders to prioritize collaborative surveillance at organizational, national, regional, or primary levels through clearly articulated and documented strategies. Another participant noted that policies support communication of the benefits of collaborative surveillance when engaging key stakeholders in collaborative surveillance. Most participants, however, cited the need to ensure enforcement, supervision, and implementation of policies would support the strengthening of collaborative surveillance systems. One participant was quoted saying:

*“Without accusing, there are policies that say, for example, every woman that attends an antenatal clinic must have syphilis testing. That’s a policy that exists in a country, and most countries achieve 80% of the people that come in readily available at antenatal clinic, it’s not 100% as well for prophylaxis prevention of ophthalmias neonatal. Yes, policies exist. But the enforcement, the supervision to assure, and ensure that policies are implemented and followed up are where the gaps are. It’s good to have a policy, but it’s another thing to have important policy and implementation*.*” MoH*

### Health Information System

Participants indicated using data was essential in prioritizing what surveillance systems should collaborate on. Participants from Malawi and Zimbabwe acknowledged using the WHONET reporting platform for AMR to report EGASP data.

Six participants mentioned collaborative surveillance using available data, and data needs to categorize priorities more effectively, coupled with resource optimization. One participant highlighted that digitization of data collection would ease data access, analysis, and use, thereby justifying collaborative surveillance to support the strengthening of systems:

“*It is easy to plan and prioritize collaborative surveillance because the data speaks better. But because all the data sets are in books, it becomes difficult to analyze them; you need to make them electronic. Imagine vaccinating more than 10 or 15 million people, and that information is in your booklet; how will you analyze it?” IP*

Another participant echoed that the first thing to do is a mindset change, understanding that the quality of data used determines the outcome product of their plan; therefore, credible plans and decisions require credible and reliable data. In support, one respondent highlighted that:

“*motivators are a difficult phenomenon to bring about and use as a driving force for enhancing surveillance because they always change. Over time, they will be pushed by activists and by interest, like political interest; now, if COVID-19 had affected more villages and confined itself to small villages in Africa, the response would not have been the same. So, motivators are defined by the people; they come and go, they’re not reliable, but data is reliable*.*” MoH*

### Health financing

To strengthen the implementation of collaborative surveillance, participants indicated that building from existing resources would be essential to pivot the new resource gaps in the system. In their descriptions, participants agreed that Zimbabwe and Malawi laboratories capacity building received support from the Fleming Fund, an entity supporting LMICs to generate, share, and use data to improve AMR surveillance through the One Health approach. Three participants stated that the funding supported more than thirty laboratories to generate AMR surveillance, including five veterinary laboratories, seven for human health, one dedicated towards food, and one for the environment across Zimbabwe and Malawi. Building on this momentum, another participant indicated that the WHO trained sixty Zimbabwean nationals from MoH, Lands, Agriculture, Fisheries, Water, Climate and Rural Development (MoLAFWRD) and Environment, Climate, Tourism and Hospitality Industry (METHI) to support the AMR surveillance. With the available resources, the EGASP funding was focused on procuring Etest strips, refresher training, and supportive visits.

Two national managers indicated having surveillance systems is costly, and an estimated budget informs making realistic decisions and areas of prioritization on collaborative surveillance systems. One participant responded that:

“*But I think, as I mentioned before, it is important that when it comes to financing, we understand exactly how much it costs us to do surveillance, what resources we need, then we’ll be able to source funds according to our need and prioritization. If we do not understand how much it costs us, we will always feel like we don’t have enough to plan collaborative surveillance*.*” FA*

Participants stated that reliance on public funding sources is the ideal funding scenario that promotes the prioritization of collaborative surveillance. All participants mentioned that collaboration is made easy when the government prioritizes surveillance funding augmented by donor funding. One participant supported this by saying:

“*I think political leaders and decision-makers tend to think that surveillance is the responsibility of a funder. But, for instance, with STIs, we’re doing syndromic management. You cannot just do central syndromic management without surveillance because you need to validate those treatment algorithms continuously, and without surveillance, it’s not going to be possible, yet that is not considered a priority for funding. The only possible way is to ride on existing funding for AMR to run the few samples we can, but that is also not sustainable*.*” MoH*.

### Health Workforce

Participants stated that investment in health workforce education and training matches the population’s needs as a driver to support collaborative surveillance. One participant stated that having a skilled workforce who understand the importance of surveillance and are keen and able to use and interpret surveillance data supports prioritizing collaborative surveillance. Some participants suggested that having a skilled workforce at all health facilities drives the plan to collaborate on surveillance systems. For EGASP, participants stated that sentinel sites were selected based on adequate staffing of health facilities to ensure program continuity. Participants stated that building a resilient and motivated workforce with retention strategies is key to implementing collaborative surveillance and systems strengthening.

One respondent indicated that:

“*If countries in the African region cannot afford to have staff specifically hired to conduct routine surveillance, it complicates the patient flow, and the current budgets do not allow, rather governments and other stakeholders should try to make the available workforce adopt the guidelines, this has been helping other countries in deploying health workers to rural, remote, and underserved areas*.*” FA*

### Service delivery

Participants agreed that providing an adequate service requires collaborative effort, support, and relationship building among staff and management. Clear communication and involvement were cited as integral to having a collaborative surveillance system with the potential to strengthen health systems. Participants indicated the need to streamline activities according to the capacity of staff and infrastructure, which could promote the implementation of collaborative surveillance where the workload and staff are balanced. One participant was quoted saying:

*“One of the most depressing terminologies that I came across, unfortunately, was task shifting and task sharing. And it meant that you are a doctor but be a clerk and a laboratory person simultaneously, and you cannot. As such, collaboration is only feasible where the workforce and the demand for service are aligned*.” *IP*

### Logistics and supplies

Four participants cited strong stakeholder coordination and inter-agency engagement, while three indicated empowering countries to prioritize local markets as essential to having the commodities and supplies. One participant, however, stated:

*“if governments have resources to support the procurement of TB and Malaria commodities, the same strategy should be channeled to procurement for all other systems; that way, implementation is made easy*.*” MoH*

Learning from EGASP, using WHO collaborating centers, and engaging with neighboring countries already implementing the same projects facilitated the procurement and shipment of supplies to the EGASP participating countries. Early procurement of commodities during the initial stage of engagements also allowed supplies to be received well on time before implementation.

#### Challenges faced in implementing a collaborative surveillance system

All participants agreed that LMICs face a myriad of challenges while implementing collaborative surveillance systems. Two national leaders cited an example of staff shortage at laboratory and sentinel sites, which threatens implementation sustainability as staff become overwhelmed with work. One public health leader, however, indicated that careful consideration of tasks is required to avoid duplicating work and roles. Another participant stated:

“*Having dedicated staff is expensive and not sustainable; countries, therefore, try to train at least everyone depending on funding, but again, with high health worker mobility, this is a huge challenge*.*” IP*

Non-availability of commodities was cited as a critical challenge to having a collaborative surveillance implementation by participants. One participant indicated that most commodities specifically for laboratory tests are manufactured out of Sub-Saharan Africa, rendering it a bottleneck in engaging collaborative surveillance; either the commodities become too expensive or unavailable, taking months before they are delivered, which heavily impacts timelines. The risk of having commodities expire on hand before use due to bureaucratic local and external approvals was cited by five participants as detrimental to collaborating systems.

Lack of support from stakeholders from government policymakers, communities, implementing partners, and other sector ministries has been cited as a significant setback in implementing collaborative surveillance. One participant called attention to the EGASP surveillance being implemented and key stakeholders not being involved. In support, an MoH participant stated that:

*“stakeholders are key in driving the program, and hence, when left out, other parts to promote collaboration will pull down efforts regardless of having all plans and resources in place. Such include other health departments like health promotion, other sector ministries, implementing partners, and civil societies and communities*.*” MoH*

Where funding is available, one manager highlighted that most of the resources go into commodities like medicines and consumables, with very little going into infrastructure upgrades or human resource capacity development, restricting capacity to do mapping of what other programs and services can be collaborated. In unison, participants cited lack of funding as a key driver in stalling collaborative surveillance. One participant stated:

*“… for example, the net budget for financing routine surveillance is donor dependent, with approximately 60% of all program specific, like the HIV and TB programs, up to 90% donor funded, the government funding is very little to none, funding emerges when there is an emergency, how then do you plan collaboration?” MoH*

Lack of accuracy, timeliness, and use of data was also a challenge to collaborative surveillance, cited by four participants. One international participant indicated that countries lack mechanisms that collect data needed to make informed decisions. At the same time, another argued that mechanisms to collect data are available, and what is lacking is the ability to analyze and use the data to inform collaboration. The same participant cited that most LMICs have become so donor-dependent that they take every program that comes with funding without looking at the country’s priorities; such programs make collaborating challenging due to the funding and mechanism restrictions even though they could be aligning. Therefore, health leaders were cited for lacking the power to negotiate for collaboration of surveillance programs.

All participants pointed out that programs tend to maintain parallel reporting systems, which creates a heavy burden for data collection, analysis, and use. One participant cited that the WHONET platform is not integrated with the national level’s District Health Information System 2 (DHIS2), hindering data sharing and access to credible data to inform collaborative surveillance.

#### Recommendations when Implementing a Collaborative Surveillance

Several recommendations for implementing collaborative surveillance were discussed by participants using their knowledge of EGASP as well as past experiences. Participants recommended using sector-wide approaches, human center design, data, existing resources, collaboration, line-ministries coordination, and existing policies when implementing collaborative surveillance systems. Key informants indicated that for health leaders to plan collaborative surveillance, there is a need to increase accountability, transparency, and responsiveness to action plans. A participant from MoH encouraged supported people-centered care approaches, stating that empowering communities in health decision-making processes will support collaborative implementation and responding to community needs. Another participant recommended that Health leaders ensure coordination and inclusivity of projects through multisectoral, multi-stakeholder, public, and private sector collaborations. Table 3 summarizes the recommendations provided by participants.

**Table 3:** Recommendations for implementing a collaborative surveillance system.

| Health systems focus | Main Challenge | Approach |
| --- | --- | --- |
| **Governance & Leadership** | No policies and enforcement of policies, if available | A people-centered approach using quality and evidence-based decisions. |
| **Health Financing** | Low or no public funds with high external funding dependence | Use a sector-wide approach to mobilize and manage resource allocation through resource mapping and needs assessments.<br>The government should specify and increase the budget allocated to surveillance. |
| **Health Information System** | Fragmented and siloed health information systems (disease- or funder-specific) are not linked to the national data warehouse systems. | One entirely digitized national health information system that can provide visual analysis for data at all health facility levels. |
| **Health Workforce** | Shortage of staff, inadequately trained workforce | Investment in education and training of health workers to match employment needs |
| **Service Delivery** | Fragmented and siloed systems | Strengthen primary health integration of services policies – not siloed systems. |
| **Medical Supplies** | Expensive commodities due to no locally available manufacturing capacities and complex supply chain systems. | Countries to leverage universities and external partnerships to manufacture their own reagents and testing kits |

## Discussion

The present study demonstrates an inseparable relationship between HSS pillars when considering collaborative surveillance learning from the EGASP implementation and evidence from the health leaders. The key to successful collaborative surveillance remains stakeholder engagement and consultations, and countries take a front-line role in steering and leading the processes. The approach can be applied at community, subnational, national, regional, and global levels, relying on shared and effective governance, communication, collaboration, and coordination. Leadership and management capacities require effective advocacy, governance, financing, resource mobilization, and quality improvement of these complex efforts to sustain health systems (21,22).

EGASP countries, together with WHO and WHO collaborating centers, have demonstrable evidence of successful collaboration by utilizing the existing standardized health systems and laboratories. To implement in the routine setting, using experiences from other public health programs implementation provides context-based decisions that can be replicable when considering other interventions of collaboration. To achieve successful and sustainable collaborations that maximize resources, private and public stakeholders can learn from other programs or interventions that are successfully collaborated (23,24). Collaborative surveillance in a routine setting involves multiple entities or organizations working together to collect, analyze, share information, or resources to enhance situational awareness, detect threats, and respond effectively (25,26). More importantly, collaboration breaks information silos, improving data flow between organizations for better decisions (27). By sharing information and resources, organizations can avoid duplicating efforts and allocate resources more effectively.

The strength that lies in good governance exhibited through accountability, transparent leadership, and collaborative routine surveillance is often underestimated for its potential to strengthen health systems. Clear, binding policies and legislation allow stakeholders to make decisions, secure resources, and collect data without constant change or fear of political persecution. Although political influence and support are essential, this often derails progress when power shifts from one hand to another. Good governance includes having policies that protect investments and steer support for technical leaders who, most often, are responsible for making decisions to ensure service integration across all health system pillars (28,29).

The results of the present study showed that collaborative surveillance requires holistic attention to how all health system pillars perform. No pillar is superior to the other, nor alone would collaborative surveillance be effective. The pillars work hand in glove, and prioritizing one and not the other potentially affects the performance and function of the other. Balancing new and existing tasks is essential for successful collaboration to ensure quality service provision and data management, which informs better decision-making (30,31). A sustainable collaborative program is guaranteed when continuous oversight of the entire system’s performance is provided. It is critical to note the link and ripple effects of lack of funding, donor dependency on the health workforce, health information systems, and supplies and commodities in strengthening collaborative surveillance systems (32). Although mobilizing local funds to support collaborative surveillance positively impacts the strengthening of systems, it is only a starting point to enhancing the performance of other health system pillars. As the world’s focus shifts to globalization and sustainable funding, governments in Africa ought to take a leaf from the after-shocks of COVID-19 and realize that if COVID-19 had only happened in Africa, the response would not have been the same (33,34).

Advocacy and digitalization are powerful tools necessary for strengthening health systems, yet very few individuals and institutions prioritize empowering health managers and technical leaders in these areas. Not all health leaders with the power to make decisions have the knowledge of data use and interpretation. To plan an effective collaborative surveillance system, investment in digitalization and focusing efforts on data capturing, analysis, use, and dissemination should be a priority for health stakeholders (35). Developing, strengthening, and reviewing health policies relies on data. With data, health managers should be adequately prepared to convince policymakers, legislators, financing agencies, and private organizations of the need to prioritize collaborative surveillance, which potentially strengthens health systems.

## Limitations

The study depended on having access to key persons coordinating, collaborating, managing, or implementing EGASP and AMR surveillance in Zimbabwe and Malawi only. Given the limited number of leaders in that area, the study relied on country visits, reports, and grey literature from the implementations to improve the validity of the results.

## Conclusions

EGASP and national AMR programs collaborated on surveillance, bringing a potential resource to providing the evidence required by countries to validate and review syndromic treatment guidelines, potentially strengthening health systems. There is a need to strengthen collaborative surveillance systems in LMCs using a people-centred approach that increases transparency, accountability and empowers national institutions and communities for sustainable activities. To achieve a sustainable and successful collaborative surveillance system that maximizes resources from both private and public sources, country-led stakeholders’ engagements are key to using contextualized evidence in decision-making when choosing the best approach to implementation. Advocacy and digitalization are essential for strengthening the surveillance systems requiring direct investments. Lessons learned from EGASP implementations in Zimbabwe and Malawi serve as models for other countries that plan to implement or improve collaborative surveillance systems for a routine setting in their context.

## Data Availability

The data is stored in a secure SharePoint folder on a computer with a protected password and is shared upon request to the lead researcher.

